# HOW IS THE IMPACT ON PUBLIC HEALTH OF SECOND WAVE OF COVID-19 PANDEMIC COMPARED TO THE FIRST WAVE? CASE STUDY OF ITALY

**DOI:** 10.1101/2020.11.16.20232389

**Authors:** Mario Coccia

**Affiliations:** CNR -- NATIONAL RESEARCH COUNCIL OF ITALY; IRCRES-CNR, Via Real Collegio, n. 30, (Collegio Carlo Alberto), 10024 - Moncalieri (TO), Italy

**Keywords:** COVID-19, SARS-CoV-2, Public health, Second wave, Epidemiology, Containment, Policy responses, Intensive Care Units

## Abstract

The main goal of this study is to compare the effects on public health of the second wave of the COVID-19 pandemic compared to first wave in society. The paper here focuses on a case study of Italy, one of the first European countries to experience a rapid increase in confirmed cases and deaths. Methodology considers daily data from February to November 2020 of the ratio of confirmed cases/total swabs, fatality rate (deaths / confirmed cases) and ratio of individuals in Intensive Care Units (ICUs) / Confirmed cases. Results reveal that the first wave of COVID-19 pandemic in Italy had a strong but declining impact on public health with the approaching of summer season and with the effects of containment measures, whereas second wave of the COVID-19 has a growing trend of confirmed cases with admission to ICUs and total deaths having a, to date, lower impact on public health compared to first wave. Although effects of the first wave of the COVID-19 pandemic on public health, policymakers have had an unrealistic optimist behavior that a new wave of COVID-19 could not hit their countries and, especially, a low organizational capacity to plan effective policy responses to cope with recurring COVID-19 pandemic crisis. This study can support vital information to design effective policy responses of crisis management to constrain current and future waves of the COVID-19 pandemic and similar epidemics in society.

## GOAL OF THE INVESTIGATION

Severe acute respiratory syndrome coronavirus 2 (SARS-CoV-2) is the strain of novel coronavirus that causes Coronavirus disease 2019 (COVID-19) with high numbers of COVID-19 related infected individuals and deaths in society (Coccia, 2020; Zhang et al., 2020). In this context, the main goal of this study is to analyze the first and second wave of the COVID-19 pandemic to compare the effects on public health in terms of confirmed cases, fatality rates and admission at Intensive Care Units. This study is important to explain the impact of COVID-19 pandemic to design effective policy responses for constraining the effects on public health and economic systems of on-going and future waves of the COVID-19 and similar epidemics.

### What is already known on these topics is based on some studies from different disciplines

Glass (2020) analyses four large countries in Europe and the USA with a proposed model and results reveal that policy responses based on limited containment measures can generate an impact of the second wave of COVID-19 pandemic on public health higher than the first one: “The results indicate that relaxations took effect in terms of increasing numbers of cases with dates ranging from early June in some countries to mid-July in other countries. For the European countries, results suggest relaxations ranging from 31% to 57% are underway and if current trends continue unchecked could lead to significant second waves that last longer than the corresponding earlier waves. In the case of the US, where the number of cases has already peaked for a second time, an extended version of the model suggests that the level of transmission may now be similar to that after the first peak”. Bontempi (2020) argues that from September 2020, Europe has to cope with the appearance of a COVID-19 second wave. The Italy situation compared with other large European countries (e.g., France, Germany, UK, and Spain) seems to show a lower impact on public health likely due to containment measures applied in the first wave of COVID-19 pandemic (cf., Atalan, 2020; Prem et al., 2020). Cacciapaglia et al. (2020) apply the *Epidemic Renormalisation Group* approach to COVID-19 pandemic, using data of the first wave, to simulate the transmission dynamics of this novel infectious disease as well as the diffusion across different European countries. Results of simulation model suggest that the peak of the second wave can be roughly between July 2020 and January 2021. In particular, the timing of the peak can be estimated considering different non-pharmaceutical measures of containment and mitigation and in addition: “The sensitivity of the second peak prognosis on the value of the infection rates gives a clear indication that social distancing measures and responsible individual behavior can have a strong effect if implemented early on” (Cacciapaglia et al., 2020). Instead, Renardy et al. (2020) apply a model based on discrete and stochastic network in a case study of Washtenaw County in Michigan (USA) to forecast the second wave of the COVID-19 pandemic. Results show that a delay of reopening does not reduce the total impact of the second peak of confirmed cases, but only delays it. However, simulations of the model reveal that a reduction of casual contacts between people can both delay and reduce the peak of the second wave of COVID-19 pandemic. Gatto et al. (2020), based on their transmission model, argue that restriction to mobility and human interactions can reduce transmission dynamics of the COVID-19 pandemic by about 45%. Other studies show that specific places have a high risk to be COVID-19 outbreaks, acting as *suprspreaders* (Chang et al., 2020). In particular, model by Chang et al. (2020), using cell phone data, predicts that a small minority of points of interest (called, POIs), such as restaurants and religious establishments, account for a large majority of infections; as a consequence, restricting maximum occupancy at each POI is more effective than uniformly reducing mobility. Moreover, higher infection rates among disadvantaged racial and socioeconomic people are due to their behavior of visiting more crowded and higher-risk places (Chang et al., 2020). In this context, countries and regions can apply timely containment and mitigation measures, such as personal protective equipment, school closing, cancellation of public/private events, restrictions on mass gatherings in public and private places, restriction on internal mobility and international travel, etc. to reduce the threats of accelerated diffusion of the waves of COVID-19 pandemic and similar viral agents in society (Petherick et al., 2020). Chu et al. (2020) also point out that mitigation measures based on social distancing and the use of facemasks seem to be effective to reduce the risk factors of transmission of the novel coronavirus. Instead, van Weert (2020) states that in the presence of a shortage of personal protective equipment, social distancing is a vital control measure to reduce the transmission dynamics of the COVID-19 pandemic in society (cf., Islam, 2020).

However, studies just mentioned are mainly based on models that generate simulations with computer experiments to predict eventual real effects of the dynamics of COVID-19 pandemic in different urban contexts. *What is hardly known in these research topics* is, using current data of COVID-19 pandemic, to explain whether the evolution of the second wave of the COVID-19 is generating an impact on public health higher or lower than first pandemic wave. The study here proposes an empirical analysis based on available data to explain the evolutionary dynamics of the second wave of COVID-19 compared to first one to design effective strategies of crisis management to cope with recurring waves of COVID-19 pandemic and future epidemics of new viral agents.

## METHODOLOGY

### 1.1 Data collection

The paper here is based on a case study of Italy that was the first large European country to experience a rapid increase in COVID-19 confirmed cases and deaths from March 2020. This study focuses on evolution of the first and second wave of COVID-19 pandemic in Italy. The end of the first wave of COVID-19 is detected here considering the minimum number of confirmed cases from February 2020 onwards, which is the 31 July 2020; after this date, confirmed cases begin to increase and this study considers the starting of the second wave of COVID-19 pandemic in Italy, i.e., 1st August 2020. In particular, this study considers data for 105 days from the starting of each wave for a comparable framework of analysis:

□ *First wave of COVID-19* from 24^th^ February, considering *N*=105 days
□ *Second wave of COVID-19* from 1^st^ August 2020 onwards, also considering *N*=105 days

In the context of first wave of COVID-19 pandemic, the containment measures of national lockdown and quarantine in Italy started on 8^th^ March 2020 and ended on 18^th^ May 2020 (Governo Italiano, 2020). In addition, Italy is located in the North hemisphere of the globe and the summer season started on 20-21 June 2020 and ended 23 September 2020, for a period of 92 days of warmer temperatures. This period is important for current study because some papers suggest that hot weather can reduce the viral infectivity of COVID-19: “high temperatures damage the virus lipid layer decreasing its stability and infection potential and may even cause virus inactivation, therefore lowering the transmission rate” (Rosario Denes et al., 2020, p. 4).

In the context of second wave of COVID-19, Italian government on 3 November 2020 applied different containment measures according to the impact of COVID-19 in regions in terms of level of admission to Intensive Care Units (ICUs) and other factors of health sector: *red* regions with full lockdown based on restrictions to individual mobility and closure of schools and public/private events; *orange* regions with a partial lockdown, and *yellow* regions in which people mainly have to wear protective mask against droplets of the coronavirus into the air and respect social distancing (cf., Chaudhry et al., 2020; Coccia, 2020f; Islam, 2020).

Data of the COVID-19 pandemic under study here are:

- daily confirmed cases
- daily deaths
- daily admission to Intensive Care Units (ICUs)
- daily swabs

Period under study is from 24 February to November 2020 and source of data is the Ministero della Salute (2020) in Italy.

### 1.2 Variables

Dynamics of the first and second wave of the COVID-19 pandemic in Italy is measured by:

▪ *Daily confirmed cases standardized =* ratio of confirmed cases (t) / swab test (t-2). The lag of about 2 days from swab test to the result of positivity to the novel coronavirus (confirmed case) is based on average time of laboratories to deliver results of the COVID-19 swab test that is roughly 1-2 days from the date of specimen pickup (LabCorp, 2020).
▪ *Daily admission to ICUs standardized =* ratio of admission to ICUs (t) / confirmed cases at (t-5). The lag of about 5 days from initial symptoms, positivity to swab test to the hospitalization and recovery in ICUs of patients is based on average time from diagnosis to hospitalization as explained by specific studies (Faes et al., 2020).
▪ *Daily Fatality rate =* ratio of deaths at (t) /confirmed cases at (t-14). The lag of about 14 days from initial symptoms to deaths is based on empirical evidence of some studies (Zhang et al., 2020).

### 1.3 Methods of statistical analysis

*Firstly*, data are analyzed with descriptive statistics, comparing arithmetic mean of measures just mentioned between first and second wave of the COVID-19 pandemic in Italy.

*Secondly*, each measure is represented in graphs comparing trends of the 1^st^ wave and 2^nd^ wave of COVID-19 pandemic, inserting the specific measure on *y*-axis (e.g., fatality rates) and temporal unit on *x*-axis given by progressive numbers, in which the number 1 indicates the starting of the pandemic wave (i.e., 24^th^ February for 1^st^ wave and 1^st^ August for 2^nd^ wave), the number two is the second day of COVID-19 pandemic wave, and so on. Moreover, the three indicators are also compared within the 1^st^ and 2^nd^ wave to have a comparative analysis of the overall evolutionary dynamics of COVID-19 pandemic (cf., Coccia and Benati, 2018).

*Thirdly*, the study explores relationships between variables with correlation analysis and test of association. This study extends the analysis with a regression model based on a linear relationship in which variables measuring the impact of the COVID-19 on public health are linear function of time (days from starting of the pandemic wave for a period of 105 days). The specification of linear relationship is given by a semi-log model:

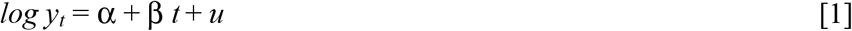

*y* = measures of the impact of COVID-19 pandemic in society: Daily fatality rate, Daily admission to ICUs, Daily confirmed cases

*t* = time given by progressive numbers representing days of the first and second wave of COVID-19 pandemic

*u =* error term

Ordinary Least Squares (OLS) method is applied for estimating the unknown parameters of linear model [1]. Statistical analyses are performed with the Statistics Software SPSS® version 26.

## RESULTS

□ Impact of the COVID-19 pandemic on public health *comparing* 1^st^ and 2^nd^ wave

First wave of COVID-19 pandemic shows from February 2020 onwards an average fatality rate of about 24%, whereas second wave of COVID-19−for the same number of 105 days from the starting in August 2020−indicates an average fatality rate of about 1.9%. Comparative analysis of the average admission to Intensive Care Units (ICUs) shows an 87.7% in the first wave and about 13% in the second one. Instead, standardized confirmed cases with swab tests show that it is about 9% in the first pandemic wave of COVID-19 and roughly 5% in the second one (Table 1). Figures 1-2-3 show the trend of variables just mentioned confirming, *ictu oculi*, that the impact of the first wave of COVID-19 pandemic in Italy on public health has been stronger than second one in the first 105 days of the evolution of this pandemic.

**Table 1.**
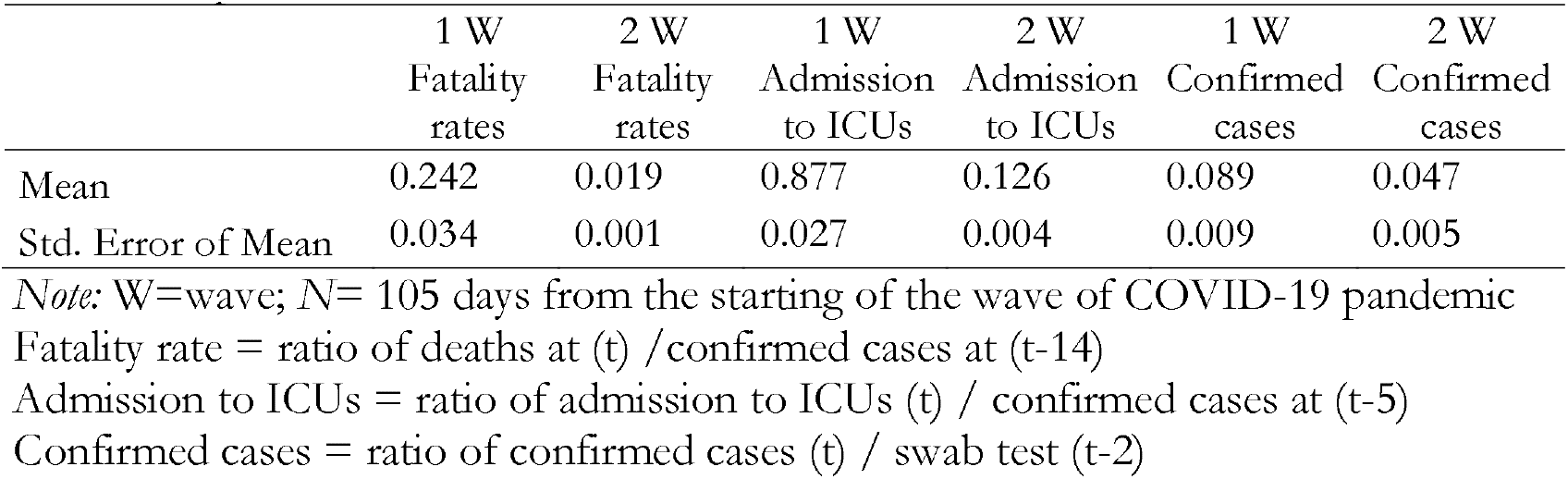
Descriptive statistics of variables measuring the impact on public health of waves of COVID-19 pandemic

**Figure 1.**
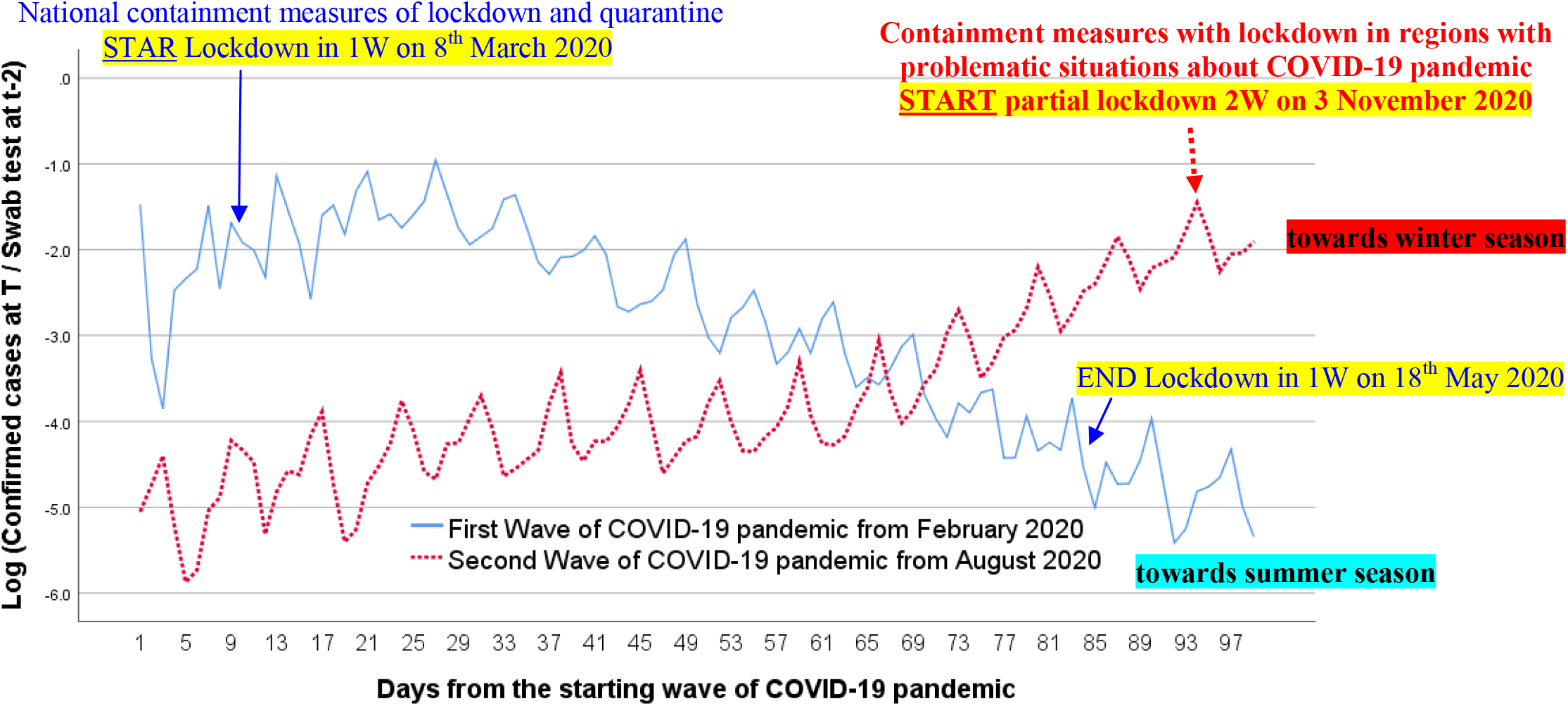
Trend of confirmed cases of the first and second wave (W) of COVID-19 pandemic in Italy, first 105 days

However, Figure 1 of confirmed cases reveals a growing trend for second pandemic wave, whereas the first one has a declining trend also because of lockdown and quarantine and the progression of COVID-19 pandemic towards summer season when the novel coronavirus seems to have a seasonality with natural reduction of transmission for better weather conditions (e.g., hot temperatures) and also low levels of air pollution for containment measures applied (cf., Coccia, 2020, 2020a, 2020b; Rosario Dentes et al., 2020).

Figure 2 shows trends of admission to ICUs: the second wave has an intensity lower than first pandemic wave and both waves seem to have stable dynamics. Instead, figure 3 shows trends of fatality rates: second pandemic wave has a low magnitude over time, suggesting a low impact on public health until November 2020.

**Figure 2.**
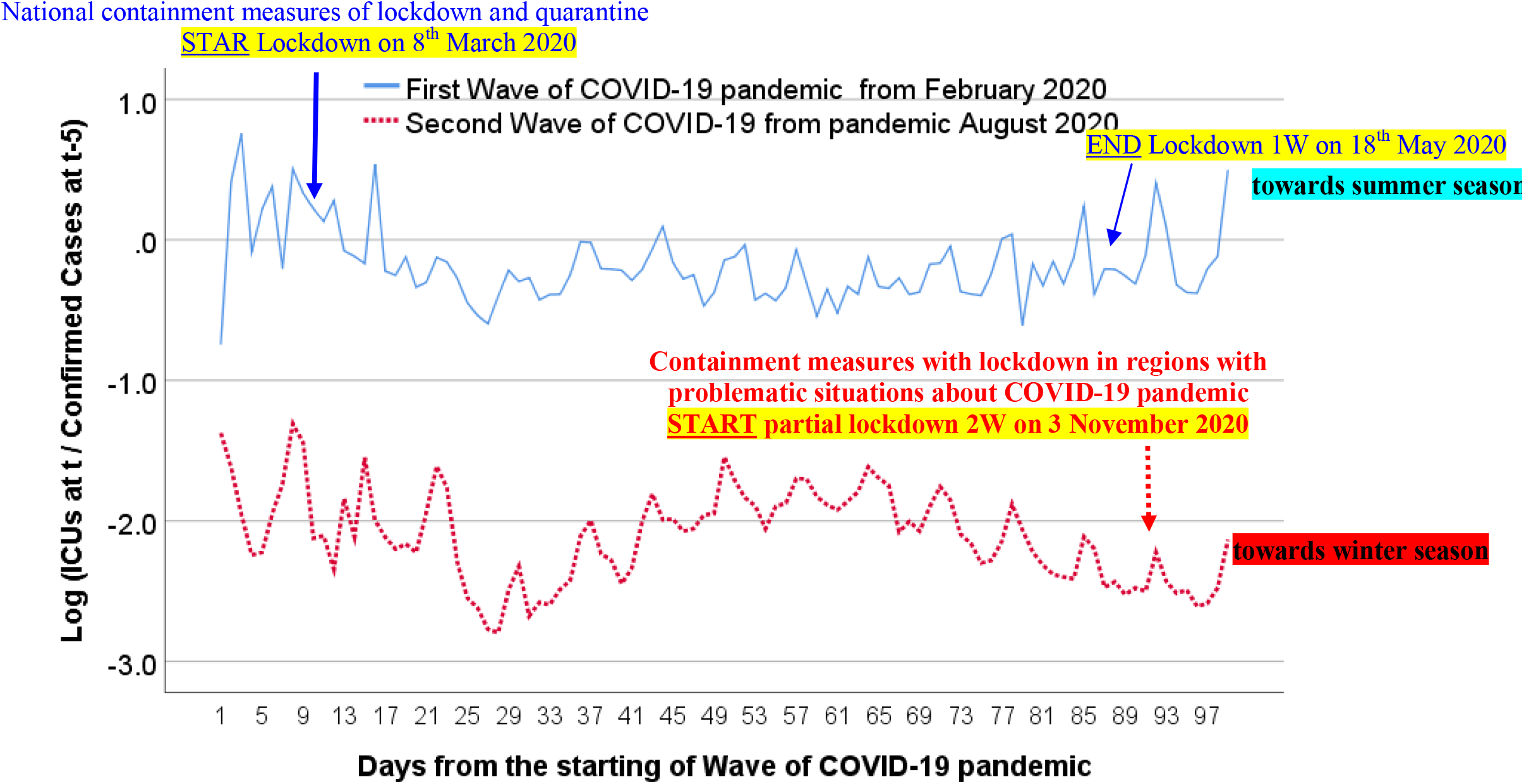
Trend of ICUs of the first and second wave (W) of COVID-19 in Italy, first 105 days

**Figure 3.**
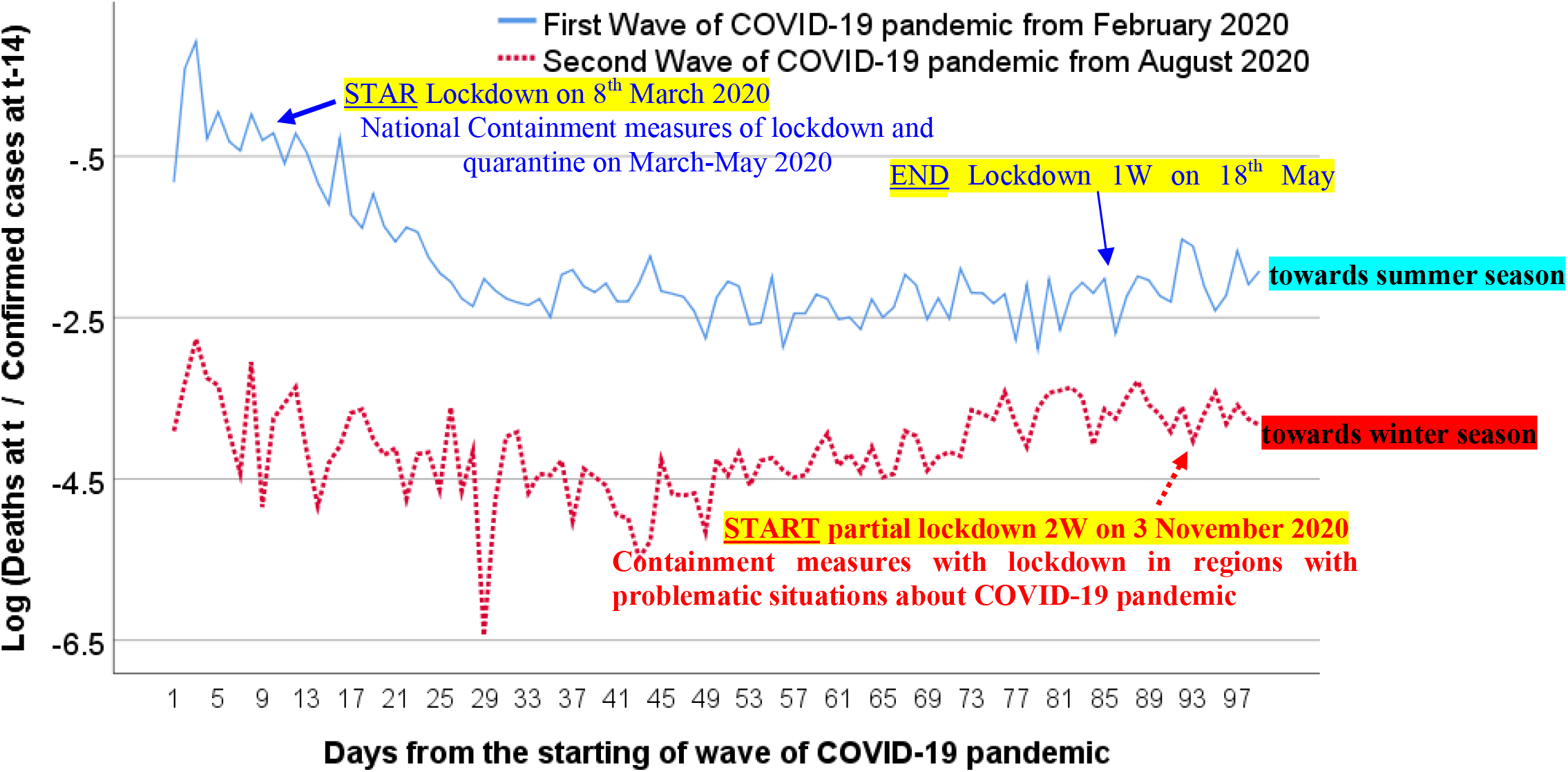
Trend of fatality rate of the first and second wave (W) of COVID-19 in Italy, first 105 days

Table 2 shows bivariate correlation analysis of variables under study in the first wave of COVID-19 pandemic: fatality rates have a high positive association with admission to ICUs (*r*=.66, *p*-value <.01), and a lower positive association of the coefficient of correlation is between fatality rates and confirmed cases (*r*=.24, *p*-value <.05), whereas correlation between ICUs and confirmed cases is negative (*r*= −.22, *p*-value <.05). Table 2 seems to show that many infected individuals died as well as a lot of patients in ICUs likely because of low knowledge of the pathology and evolution of COVID-19 in patients, and lack of appropriate therapies and low number of ICUs in hospitals (Gattinoni et al., 2020; Sterpetti, 2020). Table 3 shows tentative results for second wave of COVID-19 pandemic: correlation has a significant positive association between fatality rates and confirmed cases (*r*=.30, *p*-value <.01), whereas coefficient between ICUs and confirmed cases correlation is negative with an association higher than in the first epidemic wave (*r*=−.38, *p*-value <.01) likely because a lot of confirmed cases have not severe symptoms of COVID-19 and do not require utilization of ICUs.

**Table 2.**
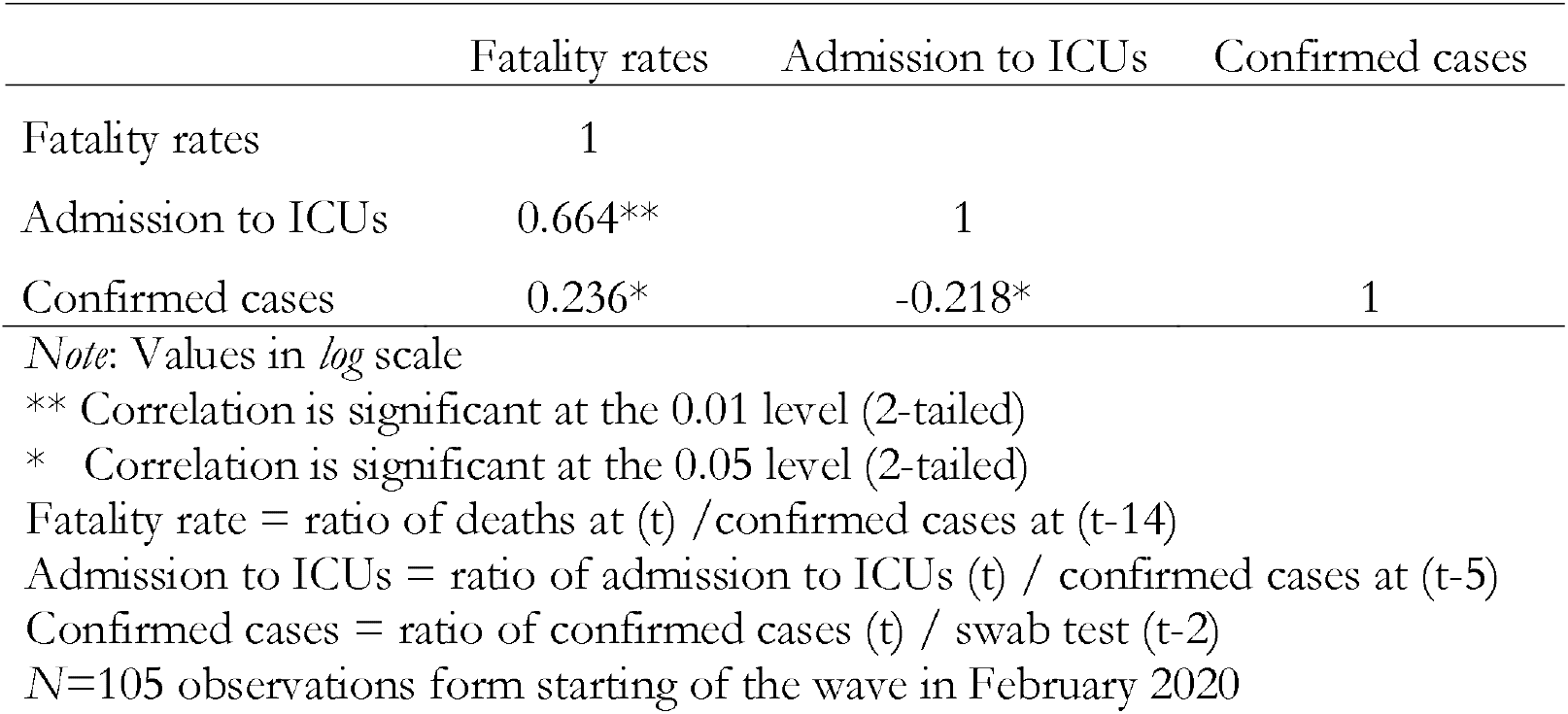
Bivariate correlation of indicators in the First Wave of COVID-19 pandemic

**Table 3.** Bivariate correlation of indicators in the Second Wave of COVID-19 pandemic

|  | Fatality rates | Admission to ICUs | Confirmed cases |
| --- | --- | --- | --- |
| Fatality rates | 1 |  |  |
| Admission to ICUs | -0.177 | 1 |  |
| Confirmed cases | 0.303** | -0.381** | 1 |

*Note:* Values in *log* scale; \*\* Correlation is significant at the 0.01 level (2-tailed).
Fatality rate = ratio of deaths at (t) / confirmed cases at (t-14)
Admission to ICUs = ratio of admission to ICUs (t) / confirmed cases at (t-5)
Confirmed cases = ratio of confirmed cases (t) / swab test (t-2)
N=105 observations form starting of the wave in August 2020

Table 4 shows the estimation of parameters in linear relationships between a number of variables and time as explanatory variable. The coefficient of regression of the model of fatality rate (dependent variable) indicates that in the first wave of COVID-19 pandemic, an increase of 1 day, it reduces the expected fatality rate by .02 (*p-value* = .001), whereas for second wave of the COVID-19, an increase of 1 day, it increases the expected fatality rate by a mere .004 (*p-value* = .05). The model’s R^2^ value indicates in the first wave that about 37% of the variation of fatality rate can be attributed (linearly) to time, whereas for second pandemic wave the coefficient of determination is rather low. The coefficient of regression of the model of admission to ICUs (dependent variable) indicates not significant results in the first wave, whereas in the second wave an increase of 1 day, it decreases the expected admission to ICUs by .003 (*p-value* = .001). Finally, the coefficient of regression using confirmed cases as dependent variable indicates that in the first wave of COVID-19 pandemic, an increase of 1 day, it reduces the confirmed cases by about .037 (*p-value* = .001), whereas for the second waves of COVID-19 pandemic, it increases by .032 (*p-value* = .001). In the last models for first and second wave of COVID-19 pandemic, R^2^ coefficient indicates that more than 76% of the variation of confirmed cases can be attributed (linearly) to time.

**Table 4.**
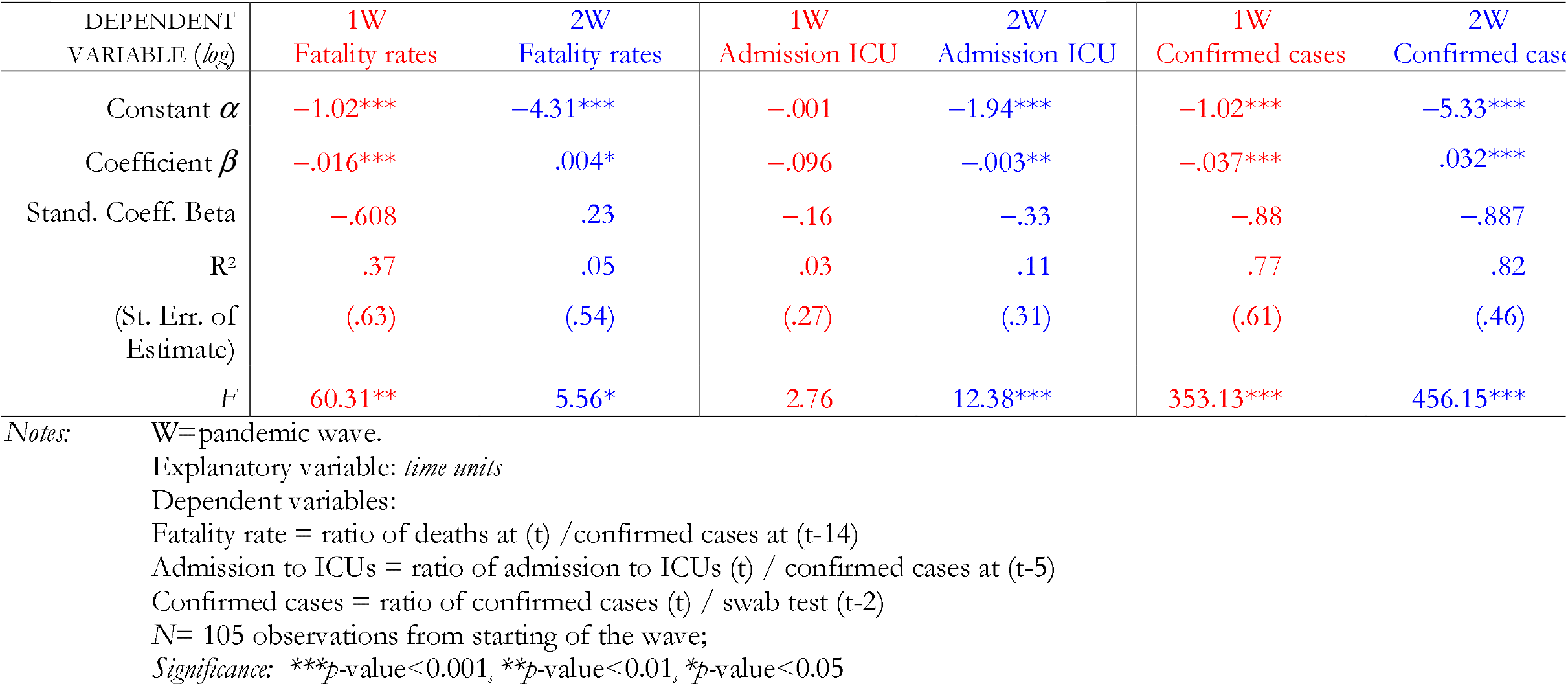
Estimated relationships, based on linear model of regression

| DEPENDENT<br>VARIABLE (*log*) | 1W<br>Fatality rates | 2W<br>Fatality rates | 1W<br>Admission ICU | 2W<br>Admission ICU | 1W<br>Confirmed cases | 2W<br>Confirmed cases |
| --- | --- | --- | --- | --- | --- | --- |
| Constant $\alpha$ | -1.02*** | -4.31*** | -.001 | -1.94*** | -1.02*** | -5.33*** |
| Coefficient $\beta$ | -.016*** | .004* | -.096 | -.003** | -.037*** | .032*** |
| Stand. Coeff. Beta | -.608 | .23 | -.16 | -.33 | -.88 | -.887 |
| R<sup>2</sup> | .37 | .05 | .03 | .11 | .77 | .82 |
| (St. Err. of<br>Estimate) | (.63) | (.54) | (.27) | (.31) | (.61) | (.46) |
| F | 60.31** | 5.56* | 2.76 | 12.38*** | 353.13*** | 456.15*** |

Notes: W=pandemic wave. Explanatory variable: *time units* Dependent variables: Fatality rate = ratio of deaths at (t) / confirmed cases at (t-14) Admission to ICUs = ratio of admission to ICUs (t) / confirmed cases at (t-5) Confirmed cases = ratio of confirmed cases (t) / swab test (t-2) N= 105 observations from starting of the wave; Significance: \*\*\* $p$ -value<0.001, \*\* $p$ -value<0.01, \* $p$ -value<0.05

General observation of regression analysis is that the first wave of the COVID-19 pandemic after national containment measures and the evolution towards summer season has a tendency to reduce fatality rates and confirmed cases, whereas during the first 105 days the second wave of COVID-19 has a low increase of fatality rate and confirmed cases but a moderate reduction of admission of patients to ICUs likely also for the evolution towards autumn-winter season when climate conditions can affect the COVID-19.

□ Analysis *within* the first and second wave of COVID-19 pandemic

In order to analyze the impact of the COVID-19 pandemic over time in society, variables under study are represented simultaneously in the same graph for 105 observations from starting day of the pandemic wave. Figure 4 shows that the first wave of COVID-19 pandemic from February 2020 has a declining trends of confirmed cases, the admission to ICUs has a high level rather stable, whereas the fatality rates after a decline in the first 30 days of the pandemic, in March or thereabout it becomes stable over time.

**Figure 4.**
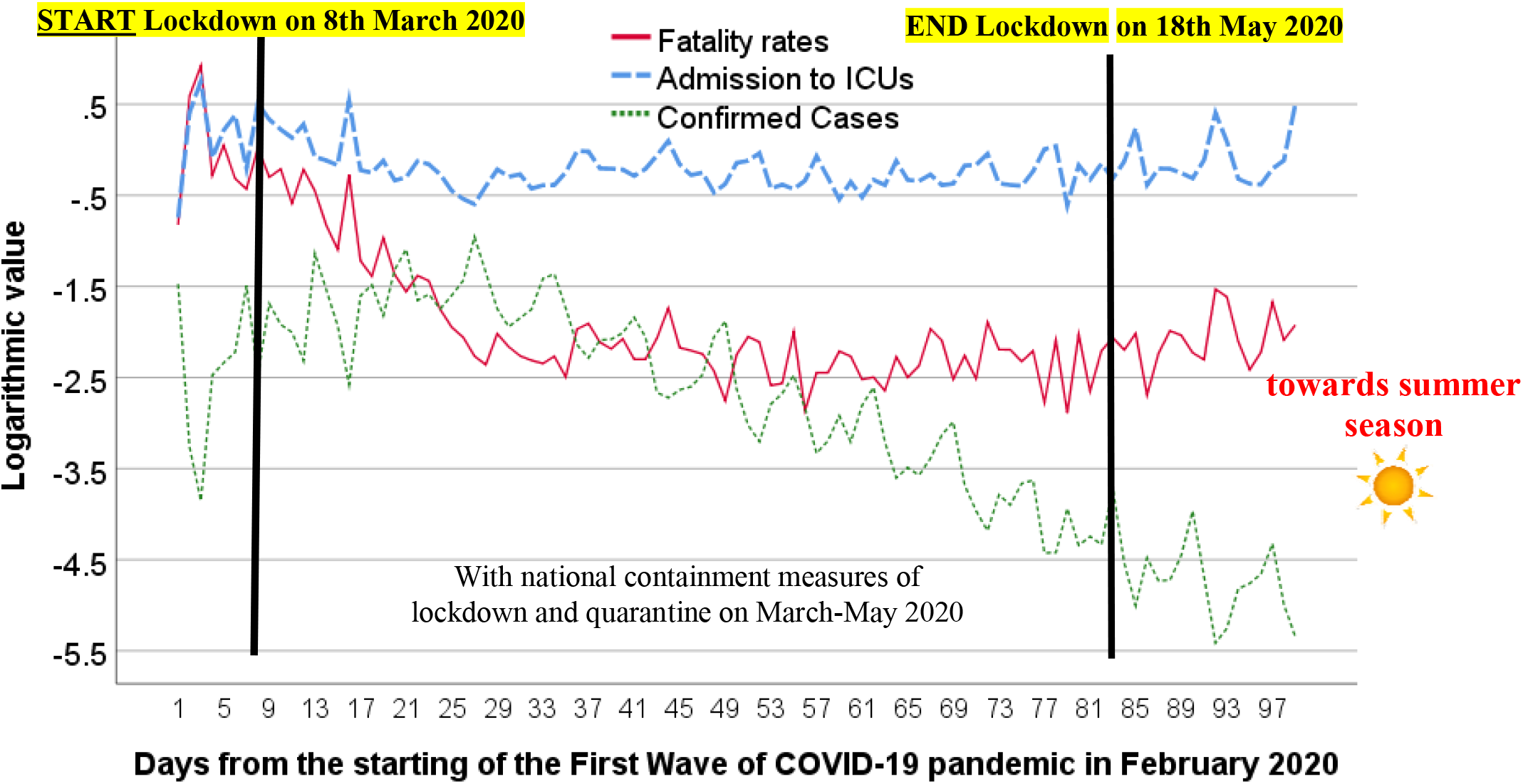
Effects of the first wave of COVID-19 pandemic on public health in Italy, first 105 days from February 2020

Figure 5 shows trends for second wave of COVID-19 from August 2020 to November 2020: admissions to ICUs are rather stable and with a level lower than first wave, whereas trend of confirmed case has a consistent grow, finally trend of fatality rates seems to have a stability in this period of autumn season.

**Figure 5.**
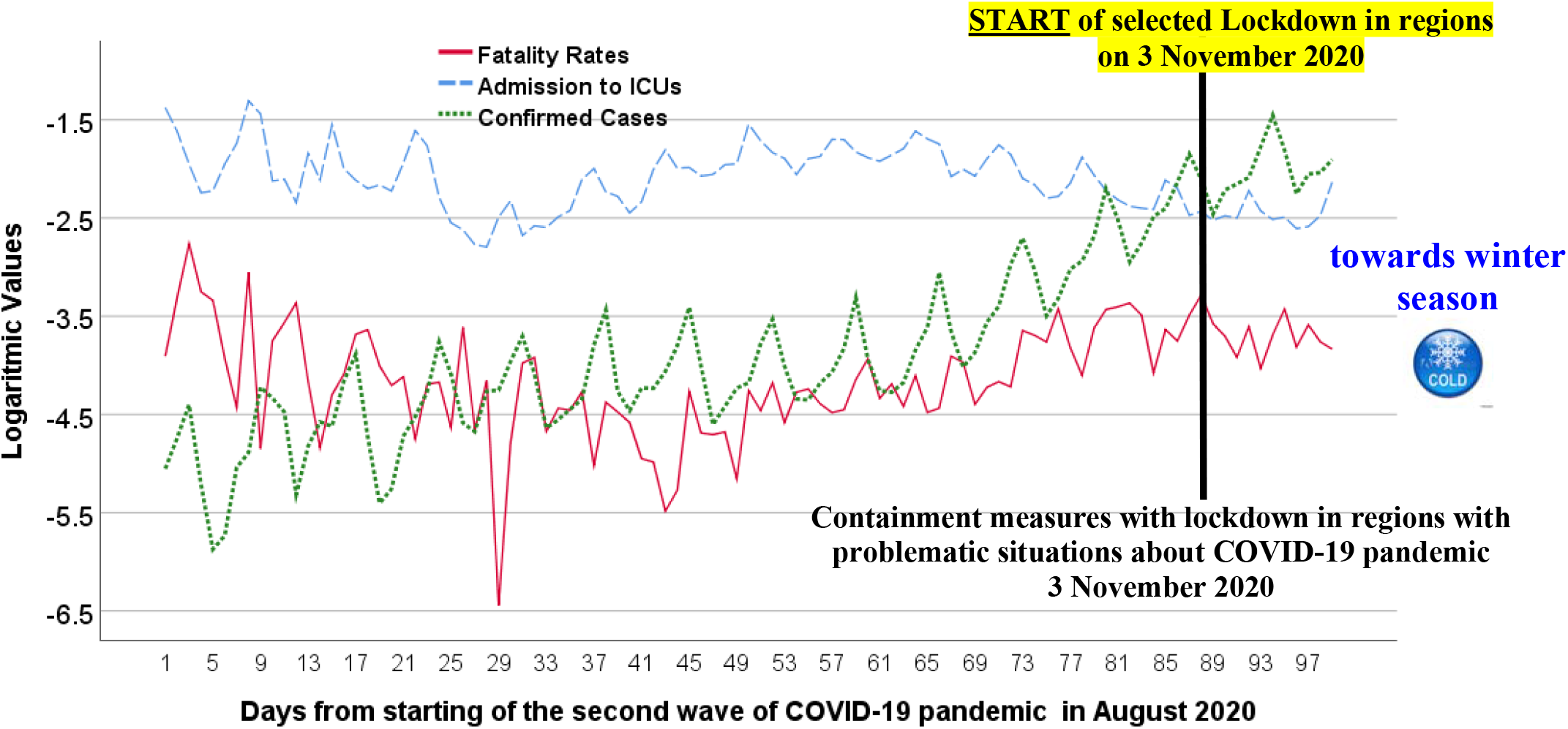
Effects of the second wave of COVID-19 pandemic on public health in Italy, first 105 days from August 2020

These results suggest that in general the first wave has had a stronger impact on public health, reduced with the approaching of summer season and national containment measures. Instead, the second wave of COVID-19 pandemic has a dynamic still in evolution that seems also to be related to climate and seasonality that may increase the impact on public health in autumn-winter season 2020-2021 like all influenza diseases, though with a lower intensity compared to the first wave of COVID-19 pandemic.

## LESSONS LEARNED

*What this study adds to current studies* on the COVID-19 global pandemic crisis is that an accurate comparison of the first and second wave of COVID-19 pandemic suggests that the first one seems to have had a stronger impact on public health, until now. In addition, government responses in the first wave of COVID-19 pandemic, based on national lockdown and quarantine, seem to have lightly constrained the diffusion of COVID-19, also helped with the approaching of summer season 2020 (cf., Coccia, 2020d, 2020f; Tobías, 2020). In general, the COVID-19 pandemic tends to have natural dynamics and seasonality that policy responses of nations seem to mitigate but without generate a significant reduction of infected cases and fatality rates (Coccia, 2020f). In fact, countries with the on-going COVID-19 pandemic have showed an uncertain governance and an unrealistic optimism about their low vulnerability that a second wave of this pandemic cannot hit them (cf., Weinstein, 1987). Although the severe impact on public health of the first wave of COVID-19 pandemic, many countries have shown still a low capability of national planning for crisis management adopting ambiguous, delayed and uncertain policy responses in the presence of recurring waves of COVID-19 pandemic crisis. In general, it seems that countries have not used in comprehensive way the process of institutional learning of the first wave of COVID-19 pandemic for supporting effective and timely critical decisions to cope with similar problematic situations generated by second pandemic wave on public health (cf., Coccia, 2018, 2019, 2020; 2020e).

## CONCLUDING OBSERVATIONS

The study here sought to understand different impact on public health of the first and second wave of the COVID-19 pandemic, analyzing a case study in Italy.

The results of analysis are:

□ First wave of COVID-19 pandemic showed an average fatality rate of 24%, whereas second wave of COVID-19 indicates an average fatality rate of about 1.9%.
□ Average admission to Intensive Care Units (ICUs) was an 87.7% in the first wave and is about 13% in the second one.
□ Average confirmed cases was about 9% in the first pandemic wave of COVID-19 and is about 5% in the second one.
□ However, confirmed cases are growing for second pandemic wave, whereas the first one had a declining trend also because of national containment measure and the progression of COVID-19 pandemic towards summer season.
□ Analysis of relationships between variables shows a high impact on public health of the first wave of COVID-19 pandemic that reduces intensity over time, whereas second wave of COVID-19 pandemic has until now a lower impact on public health but evolutionary dynamics seems to increase the intensity with the progression in the direction of winter season.

The positive side of this study is that considers a large European country, Italy, that was the first country in western world to experience a rapid increase in confirmed cases and deaths; subsequently, many countries have had a similar impact on public health of COVID-19 pandemic crisis. However, these results are based on a case study and future studies, to be reinforced in terms of generalization of suggested findings, have to enlarge the sample considering other European countries to maintain a comparable framework for statistical analyses. Hence, these conclusions are of course tentative because in the presence of the second and future waves of the COVID-19 pandemic manifold socioeconomic and environmental factors play a critical role (Coccia, 2020a, 2020b, 2020c, 2020d). There is need for much more detailed research on how COVID-19 pandemic and similar epidemics evolve in different economic, social, environmental and institutional contexts and especially in a specific period of time of a given geographical area (Coccia, 2020e). Overall, then, the investigation and explanation of the effects of pandemic waves on public health and economy are important, very important in order to design effective containment measures, apply new technologies and support R&D investments for public research directed to minimize the impact of future COVID-19 outbreaks and other epidemics similar to the COVID-19 in society, as well as interventions for not deteriorating structural indicators of the economic system of nations^1^.

To conclude, although vital results of the first wave of the COVID-19 pandemic from February to August 2020, policymakers have had an unrealistic optimist behavior that a new wave of COVID-19, started in September 2020, could not hit their countries and, especially, a low organizational capacity to plan effective policy responses to cope with recurring COVID-19 pandemic crisis (cf., Coccia, 2020f, 2020g). As a result, inappropriate and delayed policy responses associated with inefficient practices of crisis management to constrain impact of new wave of COVID-19 is again generating negative effects, *déjà vu*, on public health and of course economic systems.

## Data Availability

data are on:
Ministero della Salute 2020. Covid-19 - Situazione in Italia. http://www.salute.gov.it/portale/nuovocoronavirus/dettaglioContenutiNuovoCoronavirus.jsp?lingua=italiano&id=5351&area=nuovoCoronavirus&menu=vuoto

## Declaration of competing interest

The author declares that he has no known competing financial interests or personal relationships that could have appeared to influence the work reported in this paper. No funding was received for this study.

## Footnotes

1 ^1^ Cf., Coccia, 2016, 2017a, 2017b, 2018a, 2019a, 2020h; Forman et al., 2020.

## Notes

### Competing Interest Statement

The authors have declared no competing interest.

### Author Declarations

Not applicable', research does not report on or involve the use of any animal or human data or tissue

